# HIV seroprevalence, incidence, and viral suppression among Ugandan males with bar or sex worker partners: a population-based study

**DOI:** 10.1101/2025.03.22.25324410

**Authors:** Xinyi Feng, Kate Grabowski, Fred Nalugoda, Godfrey Kigozi, Larry W Chang, Andrea Wirtz, Caitlin E. Kennedy, Gertrude Nakigozi, Eshan U. Patel, Anthony Ndyanabo, Hadijja Nakawooya, Thomas C Quinn, Ronald M Galiwango, David Serwadda, Victor Ssempijja, Steven J Reynolds, Aaron A. R. Tobian, Robert Ssekubugu

## Abstract

**Background:** Female bar or sex workers (FBSWs) in Eastern Africa experience a high burden of HIV. However, there is limited population-level data on HIV seroprevalence, incidence, and viral suppression among their male partners.

**Methods:** Men who had sex with FBSWs in the past year were identified through longitudinal population-based HIV surveillance in southern Uganda between 2013 and 2020. Surveillance was conducted over four surveys in four Lake Victoria fishing communities (HIV seroprevalence∼40%) and 37 inland agricultural and trading communities (∼12%). Primary outcomes included laboratory-confirmed HIV seropositivity, incident infection, and viral suppression (<200 copies/mL). Prevalence and incidence rate ratios (PR, IRR) were estimated using univariable and multivariable Poisson regressions with 95% confidence intervals (95%CIs).

**Findings:** 17,438 male participants contributed 35,273 visits, with 2,420 (13.9%) reporting FBSW partners at ≥1 study visit. Men with FBSW partners tended to be older, have less education and lower incomes, and be previously married compared to those without. HIV seroprevalence was significantly higher among men with FBSW partners (vs. without FBSW partners) in both inland (21.0%vs.7.5%; PR=2.79,95%CI=2.41-3.23) and fishing communities (38.6%vs.23.0%; PR=1.67,95%CI=1.53-1.84). Overall, 154 HIV incident events occurred over 27,396 years of participant follow-up. HIV incidence was also higher among men with FBSW partners than those without (1.93vs.0.44/100 person-years; IRR=4.37,95%CI=3.04-6.16). Among men with HIV, viral suppression was similar among those with and without FBSW partners. However, the population prevalence of HIV viremia was 1.6 times higher (95%CI=1.41-1.84) among men with FBSW partners due to a higher background seroprevalence of HIV.

**Interpretation:** Men in Uganda frequently report sex with FBSWs, which is associated with a significantly higher risk of HIV acquisition. Tailored HIV prevention strategies, including the promotion and uptake of PrEP, are essential to reduce the HIV burden in this population.

**Funding:** National Institute of Allergy and Infectious Diseases, National Institutes of Health

## Introduction

Southern and eastern Africa (ESA) bears the largest burden of HIV globally, accounting for over half of the world’s population living with HIV and 37% of all new infections in 2022.^1^ Despite significant progress in reducing new infections, HIV seroprevalence remains disproportionately high among key populations, particularly female sex workers (FSWs), who face up to 30 times greater risk of acquiring HIV compared to the general population.^2^ African female bar workers (FBWs), who often engage in sex work within bars but do not self-identify as FSWs, similarly experience a high HIV burden.^3,4^ For example, HIV seroprevalence among FBWs and FSWs in Uganda is 41.6% and 31.4%,^5,6^ respectively. Qualitative studies further suggest that sex work often occurs in bar settings, yet many women prefer to identify as FBWs due to criminalization and stigma associated with sex work.^7^

While there is a growing body of research on the HIV risks faced by African female bar and sex workers (FBSWs),^8–11^ there remains a significant gap in understanding HIV acquisition and transmission risk among their male partners. The most common approach to identifying these men has been through population-based survey questionnaires, which typically ask whether respondents have paid for sex.

However, reporting payment for sex does not necessarily indicate sexual activity with FBSWs, as such exchanges may involve transactional relationships where gifts, money, or favors are exchanged for sex outside of formal sex work contexts. Less commonly, male partners have been identified through recruitment at venues or directly via FSWs or FBWs.^12–16^

A meta-analysis of 87 population-based sub-Saharan African surveys conducted between 2000 and 2020 found that approximately 8% of men reported ever paying for sex.^17^ These men were found to have a 50% higher HIV burden compared to men who did not report paying for sex. Men in central and eastern Africa were more likely to report paying for sex than those in other regions, where the relative burden of HIV among those who paid for sex was also higher. Among nine studies including data on viral suppression, no significant differences were observed between men living with HIV who did and did not pay for sex.

Studies exclusively focused on male partners of FSWs recruited through venues or via FSWs are very limited. Most of these studies were conducted prior to 2013, primarily in western Africa, and have reported a wide range of HIV seroprevalence estimates.^12^ These studies lack formal comparisons to the local male populations without FBSW partners. Furthermore, data on HIV incidence among men with FBSW partners are rare, with only one study conducted in 1987.^18^

Here, we utilized prospective, population-based data from Uganda collected between 2013 and 2020 to characterize men who reported sex with FBSWs in the past year. We then assessed HIV seroprevalence, incidence, and viremia among these men and compared it to that of the local male population not reporting sex with FBSWs. Understanding the relative HIV burden among male partners of FBSWs is critical for informing the design of targeted interventions aimed at interrupting transmission and reducing HIV burden among key and priority populations, as well as the broader African population.

## Methods

### Study design and population

Data for this study were derived from the Rakai Community Cohort Study (RCCS), an ongoing, open, population-based longitudinal HIV cohort in southern Uganda. Detailed in previous publications, RCCS conducts both household censuses and surveys in high HIV prevalence (∼40%) communities along Lake Victoria and moderate HIV prevalence (∼12%) inland agrarian and trading communities.^19^ Surveys are conducted every ∼18–24 months, following a census that records demographic and residency details.

In this study, we conducted a secondary analysis of RCCS data obtained from male participants aged 15- 49 years residing in 4 Lake Victoria fishing and 37 inland communities between July 8, 2013, and November 06, 2020. The analysis period included a total of four surveys: survey round 16 (July 8,2013- January 30,2015), 17 (February 23,2015-September 2,2016), 18 (October 3,2016-May 22,2018), and 19 (June 19,2018-November 6,2020).

### Measurement and classification of males with FBSW partners

The primary exposure of interest was self-reported sex in the past year with FSW and/or FBW partner(s). Males who engaged with FBW(s) and/or FSW(s) were aggregated due to the significant overlap between these groups shown in regional qualitative studies as well as because of their similar HIV burden.^7^ Specifically, at each study visit, men were asked about their current and past relationships within the last 12 months with up to their four most recent partners. For each partner mentioned, participants were questioned: “What are/were your partner’s main occupations?”. Participants were categorized as having sex with FBSWs in the past year at a given study visit if they reported at least one partner’s primary or secondary occupation as sex or bar workers.

### Measurements of HIV-related outcomes

The primary study outcomes included HIV seroprevalence, HIV seroconversion (i.e., incident infection), and HIV viremia. HIV serostatus was determined from participants’ venous blood samples using a validated three-test rapid HIV testing algorithm (Determine, Stat-Pak, and Uni-Gold) and confirmed through enzyme immunoassays (Murex HIV-1, 2.O).^20^ HIV viral load was measured from stored serum samples of participants with HIV using the Abbott RealTime HIV-1 Assay (Abbott Molecular, Inc., Des Plaines, IL).^21^ HIV seroconversion events were defined as instances where individuals tested HIV seropositive for the first time following a prior HIV-seronegative result, allowing for up to one missed visit. HIV viral suppression was defined as an HIV viral load of <200 copies/mL, while HIV viremia was defined as an HIV viral load of ≥200 copies/mL.

### Statistical analysis

At each study visit, we evaluated the proportion of male participants who reported having partners who were FBW, FSW, or both (FBSWs) in the past year. We summarized baseline sociodemographic characteristics and sexual behaviors of participants by frequencies and percentages, based on whether they were sexually active in the past year and ever reported having FBSW partners at any time during the study period. Pearson’s chi-squared test was used to assess statistically significant differences. For participants who reported ever having FBSW partners over the study period, the baseline visit was defined as the first visit where they reported FBSW partners. For those without FBSW partners, the baseline was considered their first visit during the study period. Additionally, we compared the characteristics of partnerships involving FBSWs and without, including the type of relationship (wife, consensual partner, girlfriend, casual friend, client/sex worker, others), duration in days, sexual behaviors (condom use, alcohol consumption before/after sex), and awareness of HIV status for both the participants and their partners.

We next assessed baseline HIV seroprevalence among men who never and ever reported FBSW partners, stratified by five-year age group and community type. We further examined HIV seroprevalence by the following: partner type (FBWs only, FSWs only, or either), number of FBSW partners among those with four sexual partners, and the length of partnership for men reporting FBSW partners only.

Next, we compared baseline HIV seroprevalence between men with and without FBSW within various demographic subgroups (e.g., five-year age group, marital status, etc.) using modified Poisson regression with robust standard errors. Associations were reported as prevalence ratios (PRs) with 95% confidence intervals (CIs), based on univariate analysis (model_1) and adjusted models for age (model_2), age and demographic variables (model_3), and age, demographic, and sexual behavioral variables (model_4).

HIV incidence was analyzed based on person-years of follow-up among participants who were initially HIV-seronegative and attended at least two consecutive survey visits (allowing one missed visit). In primary analyses, sex with FBSW partners was treated as a time-varying exposure, defined by self-reported sex with FBSW partners in the past year at the start or end of a visit interval. However, we also conducted a sensitivity analysis where the exposure was ever-reporting FBSW partners during the study period. We assumed that seroconversion occurred at the midpoint between visits. Incidence rates (IRs) were calculated as HIV seroconversions per 100 person-years. We used Poisson regression to estimate incidence rate ratios (IRRs) for HIV seroconversion, comparing men with and without FBSW partners. We conducted both unadjusted and age-adjusted analyses. We also conducted subgroup analyses stratified by community type and calendar time (rounds 16–17, rounds 18–19) and among sexually active participants only. We applied stabilized inverse-probability-of-censoring weighting to address potential selection bias from differential loss to follow-up (**Supplementary_Statistical_Method**).

Among HIV seropositive visits, we assessed viral suppression at each survey round and across all surveys by sex with FBSWs and calculated the ratios of viral suppression adjusting for age and stratifying by community type. Additionally, among all participants, we calculated the population prevalence of HIV viremia by engagement with FBSW partners at each survey round and assessed the ratios of HIV viremia at each survey round and overall surveys adjusting for age and stratifying by community type. For estimates across all survey rounds, we applied Poisson regression models with generalized estimating equations (GEE), using an exchangeable correlation structure and robust standard errors to account for within-subject correlations over time.

Among HIV seropositive visits, we evaluated viral suppression at each survey round and across all surveys based on sex with FBSWs, calculating unadjusted and age-adjusted suppression ratios stratified by community type. Additionally, among all participants, we estimated the population prevalence of HIV viremia by engagement with FBSWs at each survey round and calculated prevalence ratios of HIV viremia both at each survey round and across all surveys, adjusting for age and stratifying by community type. For estimates across all survey rounds, we used Poisson regression models with generalized estimating equations (GEE) using an exchangeable correlation structure and robust standard errors to account for within-subject correlations over time.

In the final survey round 19, the only round during which these data was collected, we evaluated HIV serostatus awareness among men living with HIV, and both the awareness and usage of pre-exposure prophylaxis (PrEP) among HIV-seronegative men, by their engagement with FBSWs. Results were summarized as frequencies and percentages, with Pearson’s chi-squared tests used to identify differences.

All statistical analyses were performed using Stata 15.1 (Stata Corp, College Station, TX) and R 4.2.1 (R Core Team, 2022). We used two-sided p-values with a threshold of <0.05 to determine statistical significance.

### Ethics

This study was reviewed and approved by the Uganda Virus Research Institute Research and Ethics Committee and the Johns Hopkins School of Medicine Institutional Review Board. All participants provided written informed consent.

## Results

### Study population characteristics

Among 26,871 eligible males aged 15-49 residing in RCCS communities, 9,344 (34.8%) were away for work or school at the time of the survey and excluded. Of those present in the community at the time of the survey, 51 (0.3%) refused study participation, and 17,476 (99.7%) were enrolled. Among these men, 17,438 (99.8%) provided a blood sample and had a valid HIV test result, contributing to 35,273 person visits to the analysis(**Supplementary_Table_1,Supplementary_Figure_1**).

Overall, 2,420 (13.9%) male participants self-reported having FBSW partners during one or more study visits. A higher proportion of males in fishing communities than inland communities reported having FBSW partners at baseline (37.1%vs.6.2%) and at each survey round (**Supplementary_Table_2**). At baseline, sexually active men with FBSW partners in the past year were typically older, more likely to live in fishing communities, had lower education and SES, more likely to be previously married, less likely to be circumcised, and more likely to have GUD in the past year. They also reported higher engagement in HIV-associated sexual behaviors (e.g., greater number of sexual partners in the past year, alcohol use before sex) compared to men without FBSW partners or those not sexually active in the past year (**Table_1**). Of 3,479 person visits contributed by men reporting at least one FBSW partner, 2,692 (77.0%) also involved non-FBSW partners.

**Table 1.**
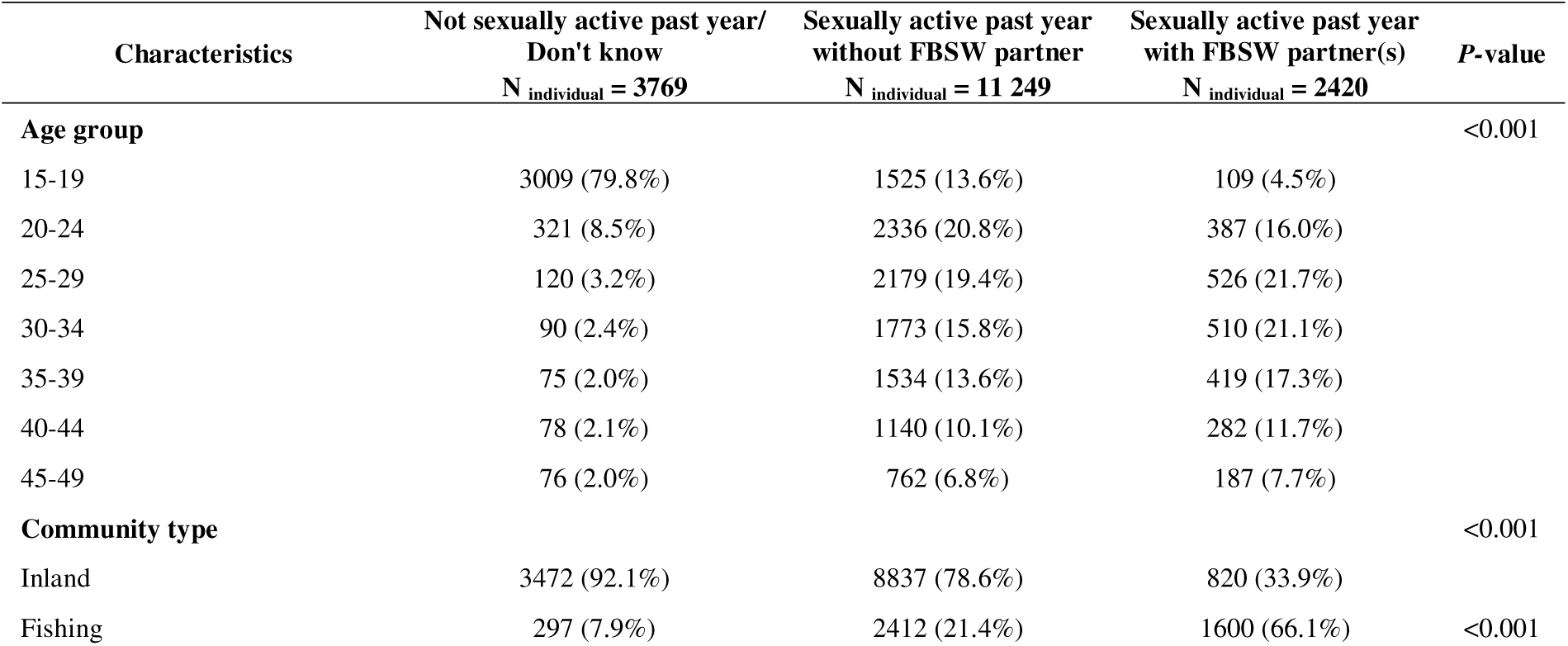
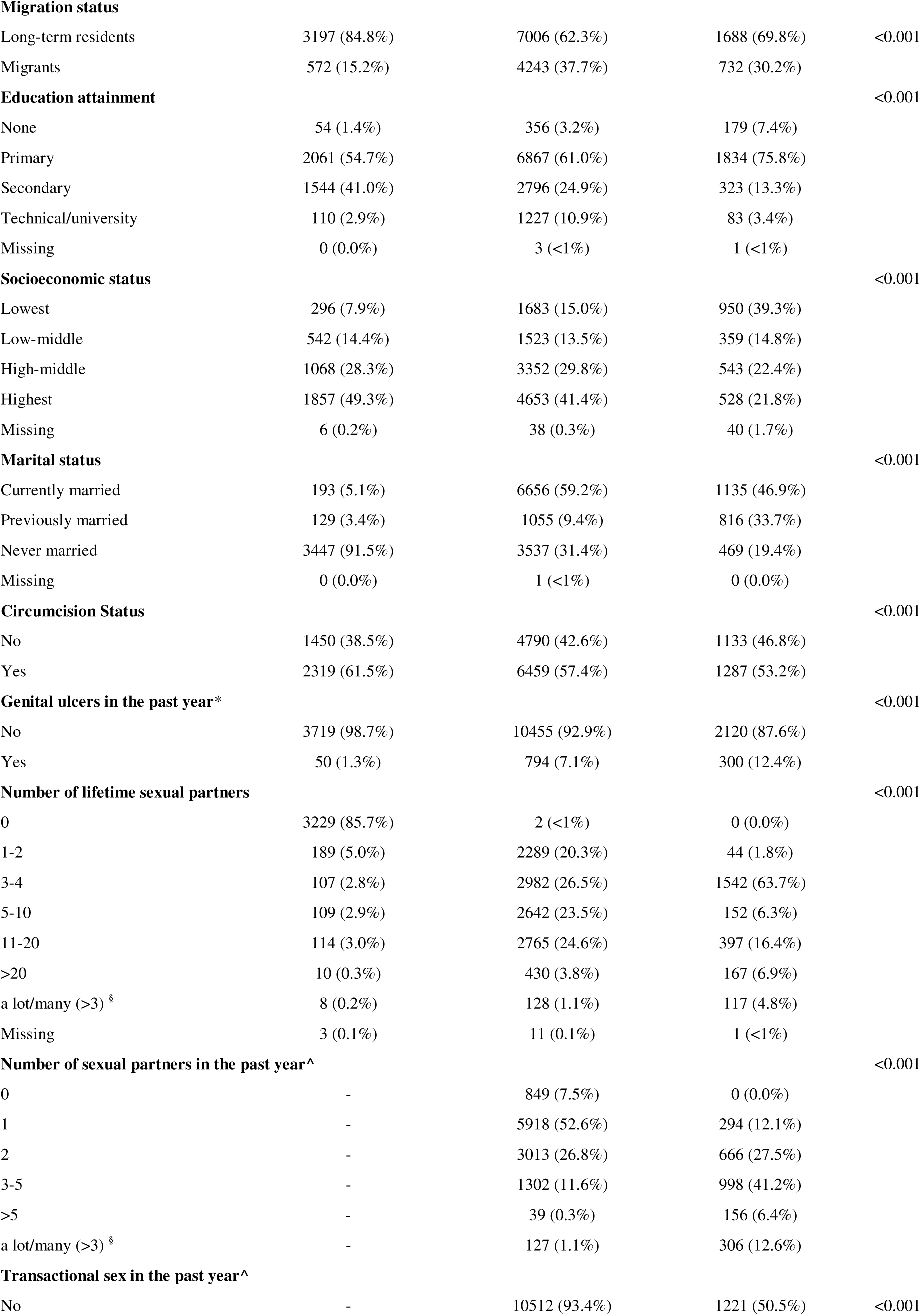
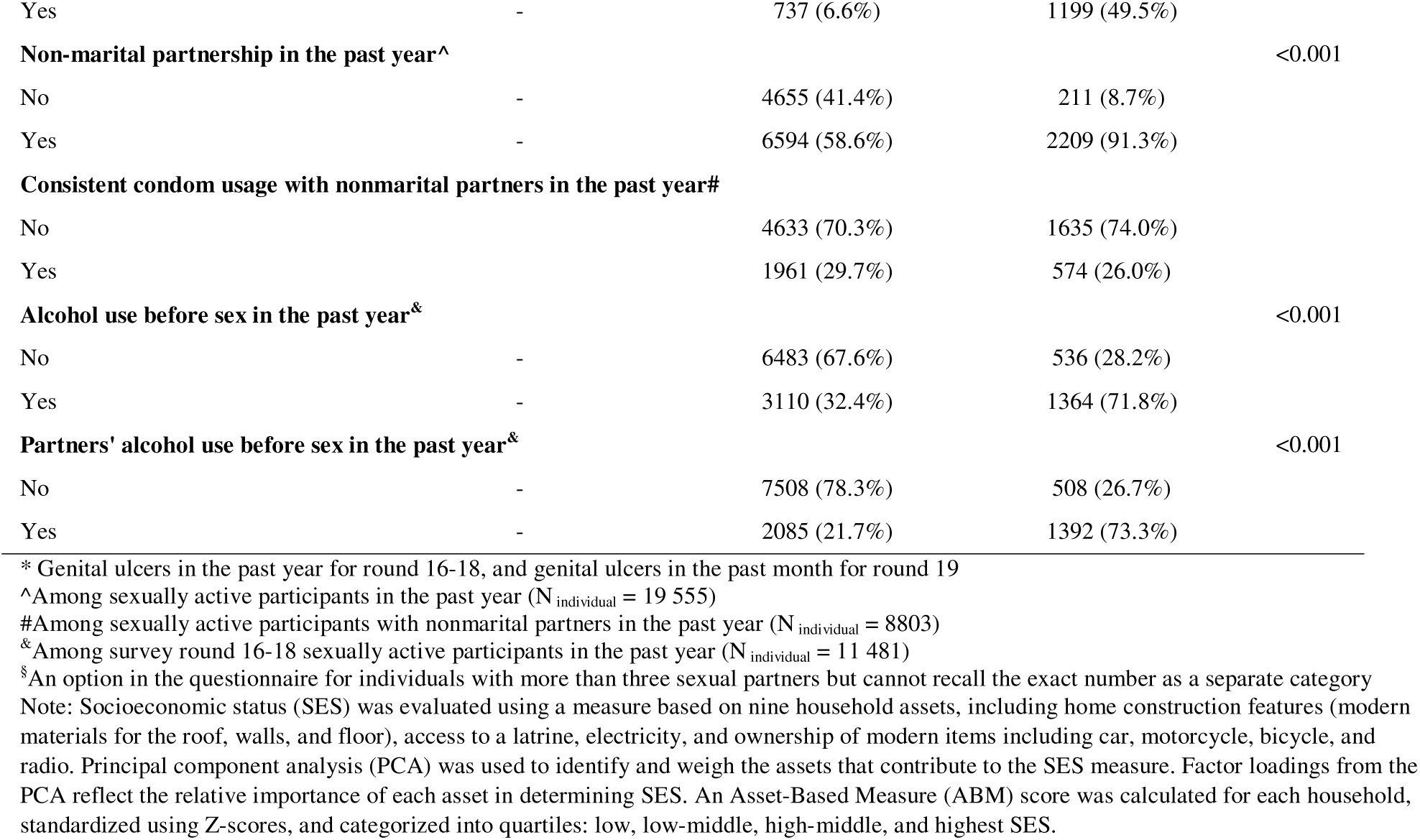
Baseline characteristics by self-reported engagement with female bar or sex worker (FBSW) partners among male participants in the Rakai Community Cohort Study, Uganda, 2013-2020 (N_individual_ = 17 438)

### Characteristics of partnerships involving FBSWs

A total of 51,437 sexual partnerships were reported, of which 5,505 (10.7%) involved FBSWs. Compared to non-FBSW partnerships, these partnerships were often one-time encounters (median length=0 days,IQR:0-90) and less likely to be ongoing at the time of the survey. Condom use was higher in partnerships with FBSWs (48.5%vs.20.2%), but 78.3% occurred without knowing the FBSW’s HIV serostatus, and 69.4% involved no disclosure of the male participant’s HIV serostatus(**Table_2**).

**Table 2.** Characteristics of sexual partnership by self-reported engagement with female bar or sex worker (FBSW) partners among male participants in the Rakai Community Cohort Study, Uganda, 2013-2020 (N_individual_ = 14 745)

| Partnership characteristics | Partnership with non-FBSW | Partnership with FBSW | P-value |
| --- | --- | --- | --- |
|  | N<sub>partnership</sub> = 45 932 | N<sub>partnership</sub> = 5505 |  |
| Length of relationship in days, median (IQR) | 365.0 (90.0, 2186.0) | 0.0 (0.0, 90.0) | <0.001 |
| Relationship type |  |  |  |
| Current wife | 6529 (14.2%) | 83 (1.5%) | <0.001 |
| Current consensual partner | 14 796 (32.2%) | 488 (8.9%) |  |
| Girlfriend | 19 876 (43.3%) | 1513 (27.5%) |  |
| Occasional or casual friend | 4046 (8.8%) | 758 (13.8%) |  |
| Client/sex worker | 224 (0.5%) | 2617 (47.5%) |  |
| Others | 461 (1.0%) | 46 (0.8%) |  |
| Relationship ongoing |  |  |  |
| Yes | 31 804 (69.2%) | 1210 (22.0%) | <0.001 |
| No | 14 072 (30.6%) | 4280 (77.7%) |  |
| Do not know | 56 (0.1%) | 15 (0.3%) |  |
| **Condom use** |  |  |  |
| Never | 27 555 (60.0%) | 2245 (40.8%) | <0.001 |
| Inconsistent | 9096 (19.8%) | 586 (10.6%) |  |
| Always | 9263 (20.2%) | 2669 (48.5%) |  |
| Do not know | 18 (<1%) | 5 (0.1%) |  |
| **Partner's HIV status was ever known by RCCS participant** |  |  |  |
| Yes | 30 340 (66.1%) | 1195 (21.7%) | <0.001 |
| No | 15 588 (33.9%) | 4310 (78.3%) |  |
| Do not know | 4 (<1%) | 0 (0.0%) |  |
| **Informed partner(s) HIV status of RCCS participants** |  |  |  |
| Yes | 20 508 (44.6%) | 1168 (21.2%) | <0.001 |
| No | 11 732 (25.5%) | 3822 (69.4%) |  |
| Never tested/got HIV results | 2417 (5.3%) | 295 (5.4%) |  |
| Received couple counseling | 11 270 (24.5%) | 220 (4.0%) |  |
| Do not remember | 5 (<1%) | 0 (0.0%) |  |
| **Uses alcohol before sex with partner(s)\*** |  |  |  |
|  | **N<sub>partnership</sub> = 33 891** | **N<sub>partnership</sub> = 4022** |  |
| Yes | 11 570 (34.1%) | 2469 (61.4%) | <0.001 |
| No | 22 321 (65.9%) | 1553 (38.6%) |  |
| **Partner uses alcohol before sex\*** |  |  |  |
|  | **N<sub>partnership</sub> = 33 891** | **N<sub>partnership</sub> = 4022** |  |
| Yes | 6825 (20.1%) | 2265 (56.3%) | <0.001 |
| No | 27 062 (79.9%) | 1757 (43.7%) |  |
| Do not know | 4 (<1%) | 0 (0.0%) |  |

\*Among male participants in the Rakai Community Cohort Study, Uganda, 2013-2018 (N<sub>individual</sub> = 12 511)
Abbreviations: FBSW, female bar or sex worker; IQR, interquartile range; RCCS, Rakai Community Cohort Study; HIV, human immunodeficiency virus

Notably, the characteristics of partnerships with FBWs and FSWs differed. Partnerships with FSWs were generally shorter, non-ongoing, involved more frequent condom use, and had lower awareness of the partner’s HIV status compared to partnerships with FBWs. Additionally, 86.3% of partnerships with FSWs were classified as client/sex worker relationships, compared to 23.9% of FBW partnerships, which were most commonly described as girlfriends (42.4%)(**Supplementary_Table_3**).

### Population prevalence of HIV

At baseline, HIV seroprevalence was significantly higher among men with FBSW partners (32.6%[789/2,420]) compared to those without (10.3%[1,549/15,018])(age-adjusted PR=2.46, 95% CI=2.28–2.65)(**Supplementary_Table_4**). These differences between men with and without FBSW partners were consistent across age groups and community types, with inland communities showing a prevalence of 21.0% versus 7.5% (PR=2.79,95%CI=2.41-3.23) and fishing communities showing a prevalence of 38.6% versus 23.0% (PR=1.67,95%CI=1.53-1.84)(**Figure_1A**). Similar disparities were observed among men across a wide range of sociodemographic profiles(**Table_3**).

**Figure 1.**
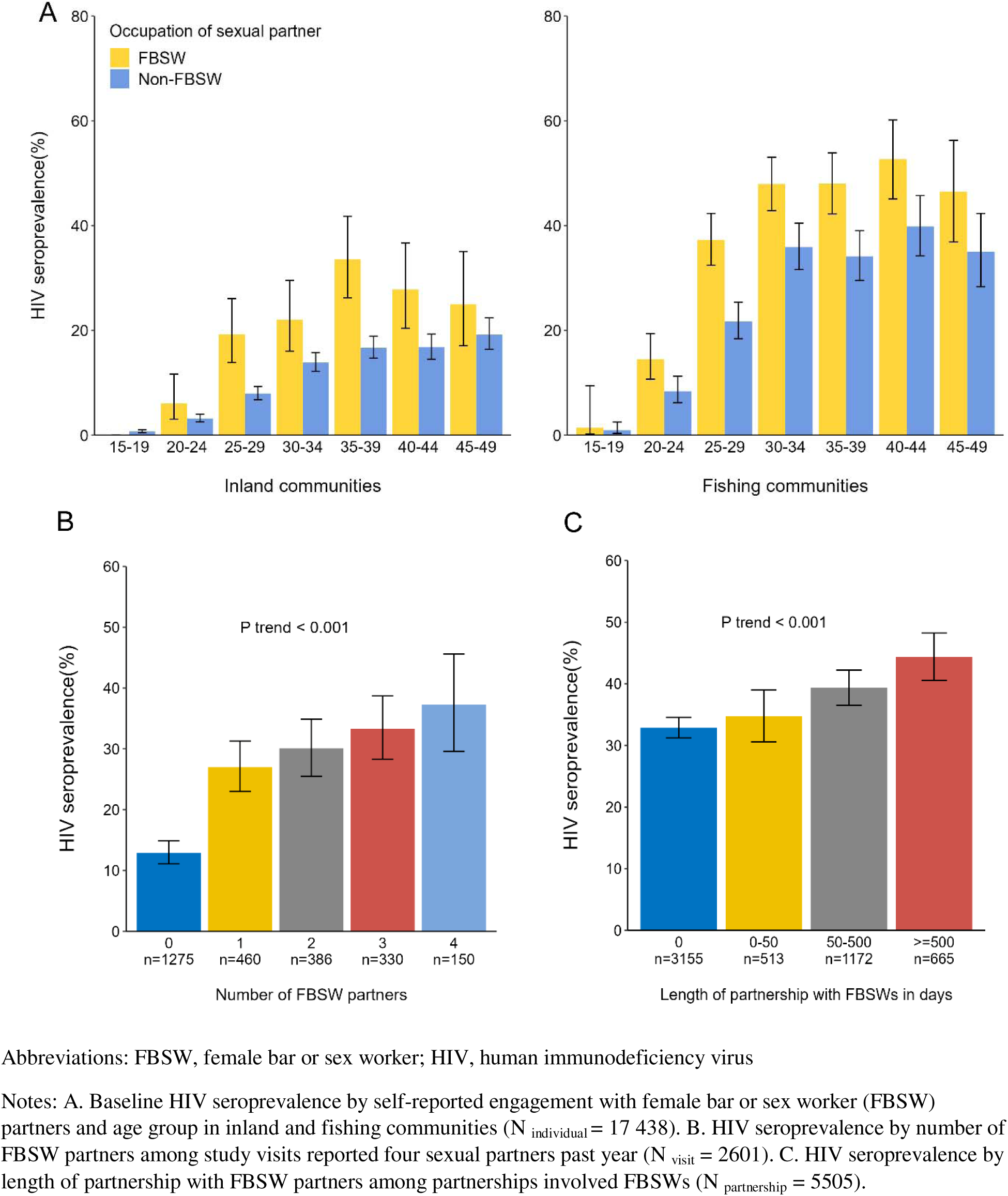
HIV seroprevalence by age group and community type, number of FBSW partners, and length of partnership with FBSW partners among male participants of the Rakai Community Cohort Study, Uganda, 2013-2020.

**Table 3.**
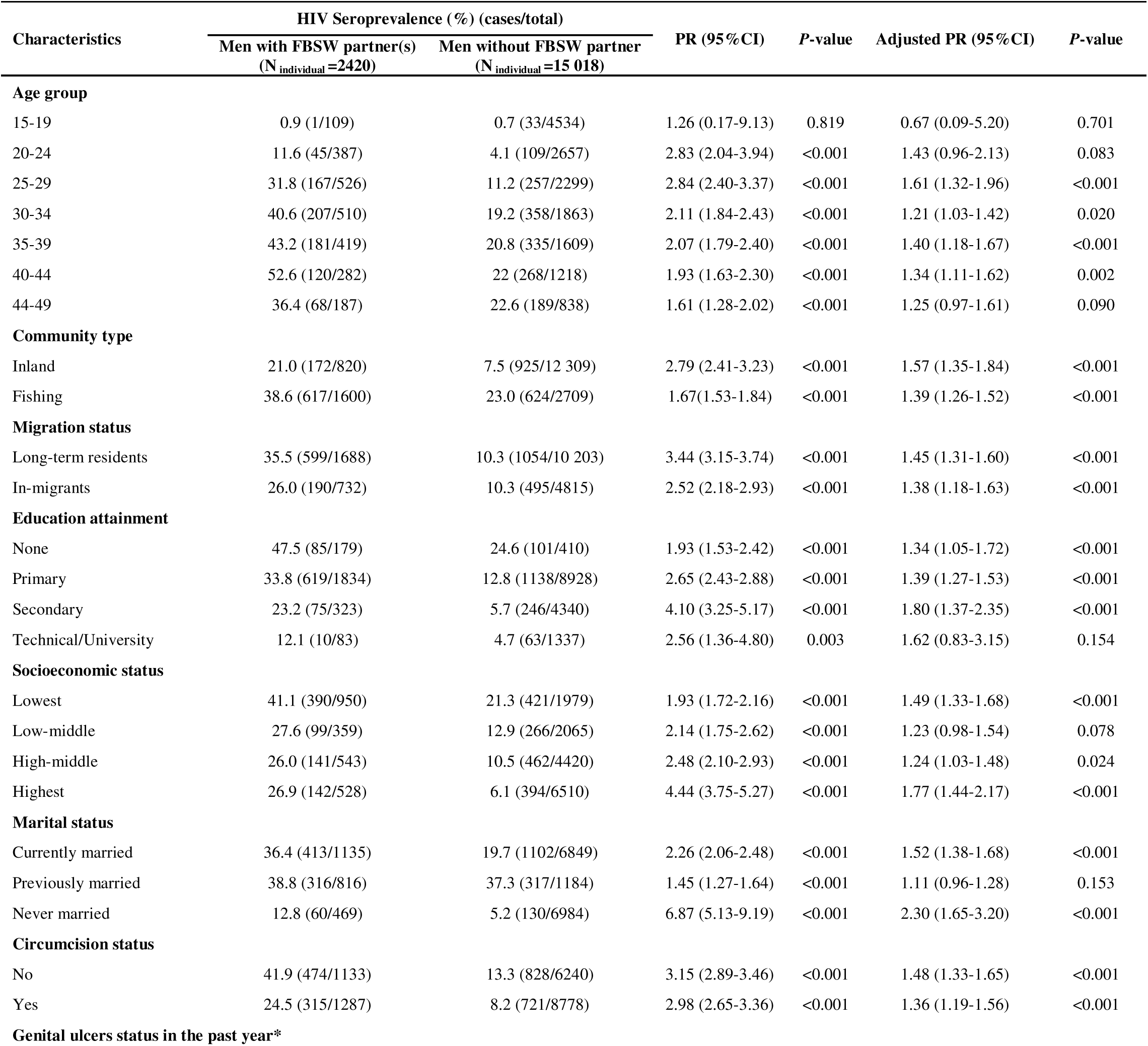
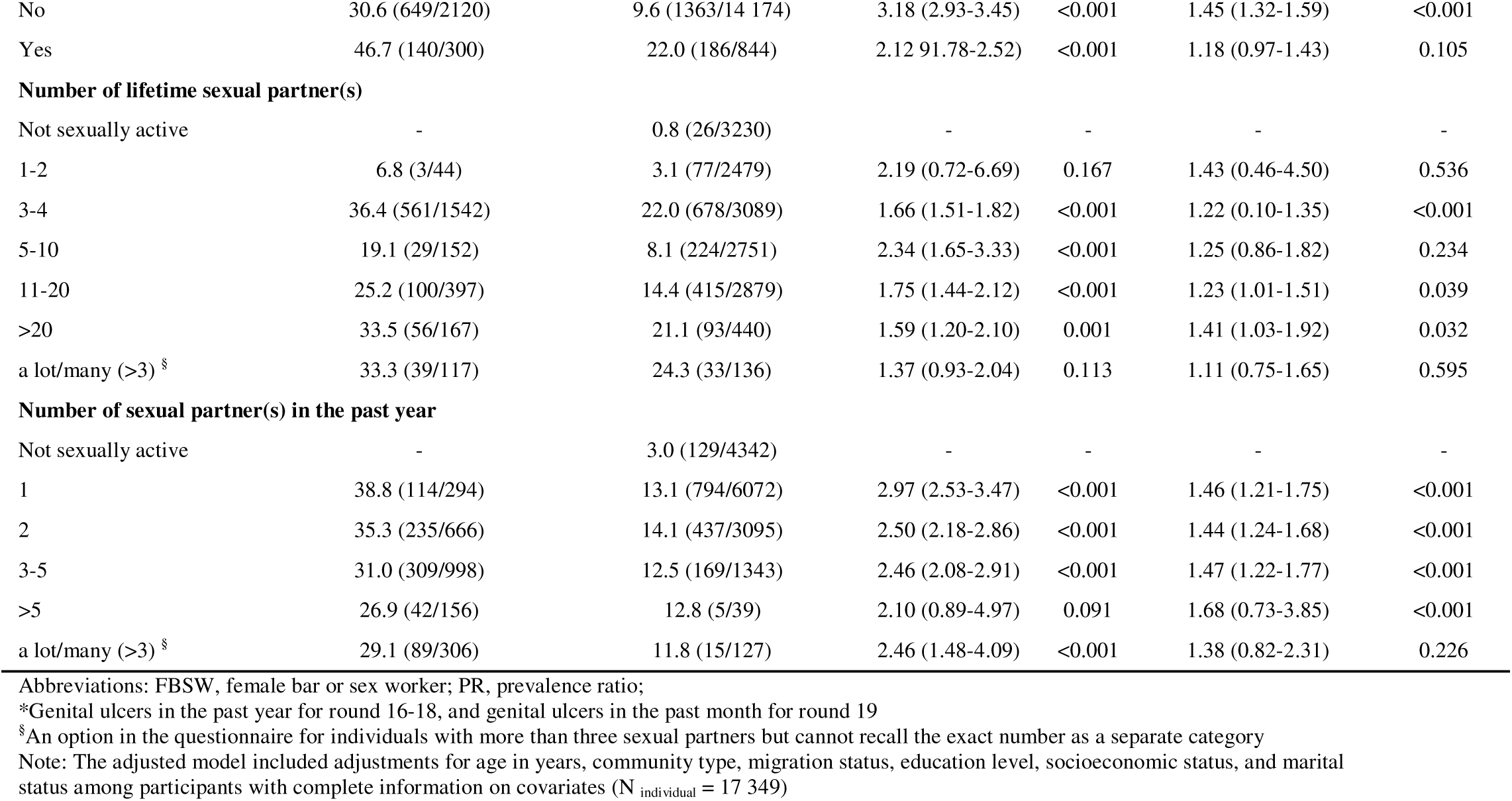
HIV seroprevalences and prevalence ratios at baseline by self-reported engagement with female bar or sex worker (FBSW partners among subgroups of male participants in the Rakai Community Cohort Study, Uganda, 2013-2020 (N_individual_ = 17 438)

| Characteristics | HIV Seroprevalence (%) (cases/total) |  | PR (95%CI) | P-value | Adjusted PR (95%CI) | P-value |
| --- | --- | --- | --- | --- | --- | --- |
|  | Men with FBSW partner(s)<br>(N<sub>individual</sub> =2420) | Men without FBSW partner<br>(N<sub>individual</sub> =15 018) |  |  |  |  |
| Age group |  |  |  |  |  |  |
| 15-19 | 0.9 (1/109) | 0.7 (33/4534) | 1.26 (0.17-9.13) | 0.819 | 0.67 (0.09-5.20) | 0.701 |
| 20-24 | 11.6 (45/387) | 4.1 (109/2657) | 2.83 (2.04-3.94) | <0.001 | 1.43 (0.96-2.13) | 0.083 |
| 25-29 | 31.8 (167/526) | 11.2 (257/2299) | 2.84 (2.40-3.37) | <0.001 | 1.61 (1.32-1.96) | <0.001 |
| 30-34 | 40.6 (207/510) | 19.2 (358/1863) | 2.11 (1.84-2.43) | <0.001 | 1.21 (1.03-1.42) | 0.020 |
| 35-39 | 43.2 (181/419) | 20.8 (335/1609) | 2.07 (1.79-2.40) | <0.001 | 1.40 (1.18-1.67) | <0.001 |
| 40-44 | 52.6 (120/282) | 22 (268/1218) | 1.93 (1.63-2.30) | <0.001 | 1.34 (1.11-1.62) | 0.002 |
| 44-49 | 36.4 (68/187) | 22.6 (189/838) | 1.61 (1.28-2.02) | <0.001 | 1.25 (0.97-1.61) | 0.090 |
| Community type |  |  |  |  |  |  |
| Inland | 21.0 (172/820) | 7.5 (925/12 309) | 2.79 (2.41-3.23) | <0.001 | 1.57 (1.35-1.84) | <0.001 |
| Fishing | 38.6 (617/1600) | 23.0 (624/2709) | 1.67(1.53-1.84) | <0.001 | 1.39 (1.26-1.52) | <0.001 |
| Migration status |  |  |  |  |  |  |
| Long-term residents | 35.5 (599/1688) | 10.3 (1054/10 203) | 3.44 (3.15-3.74) | <0.001 | 1.45 (1.31-1.60) | <0.001 |
| In-migrants | 26.0 (190/732) | 10.3 (495/4815) | 2.52 (2.18-2.93) | <0.001 | 1.38 (1.18-1.63) | <0.001 |
| Education attainment |  |  |  |  |  |  |
| None | 47.5 (85/179) | 24.6 (101/410) | 1.93 (1.53-2.42) | <0.001 | 1.34 (1.05-1.72) | <0.001 |
| Primary | 33.8 (619/1834) | 12.8 (1138/8928) | 2.65 (2.43-2.88) | <0.001 | 1.39 (1.27-1.53) | <0.001 |
| Secondary | 23.2 (75/323) | 5.7 (246/4340) | 4.10 (3.25-5.17) | <0.001 | 1.80 (1.37-2.35) | <0.001 |
| Technical/University | 12.1 (10/83) | 4.7 (63/1337) | 2.56 (1.36-4.80) | 0.003 | 1.62 (0.83-3.15) | 0.154 |
| Socioeconomic status |  |  |  |  |  |  |
| Lowest | 41.1 (390/950) | 21.3 (421/1979) | 1.93 (1.72-2.16) | <0.001 | 1.49 (1.33-1.68) | <0.001 |
| Low-middle | 27.6 (99/359) | 12.9 (266/2065) | 2.14 (1.75-2.62) | <0.001 | 1.23 (0.98-1.54) | 0.078 |
| High-middle | 26.0 (141/543) | 10.5 (462/4420) | 2.48 (2.10-2.93) | <0.001 | 1.24 (1.03-1.48) | 0.024 |
| Highest | 26.9 (142/528) | 6.1 (394/6510) | 4.44 (3.75-5.27) | <0.001 | 1.77 (1.44-2.17) | <0.001 |
| Marital status |  |  |  |  |  |  |
| Currently married | 36.4 (413/1135) | 19.7 (1102/6849) | 2.26 (2.06-2.48) | <0.001 | 1.52 (1.38-1.68) | <0.001 |
| Previously married | 38.8 (316/816) | 37.3 (317/1184) | 1.45 (1.27-1.64) | <0.001 | 1.11 (0.96-1.28) | 0.153 |
| Never married | 12.8 (60/469) | 5.2 (130/6984) | 6.87 (5.13-9.19) | <0.001 | 2.30 (1.65-3.20) | <0.001 |
| Circumcision status |  |  |  |  |  |  |
| No | 41.9 (474/1133) | 13.3 (828/6240) | 3.15 (2.89-3.46) | <0.001 | 1.48 (1.33-1.65) | <0.001 |
| Yes | 24.5 (315/1287) | 8.2 (721/8778) | 2.98 (2.65-3.36) | <0.001 | 1.36 (1.19-1.56) | <0.001 |
| Genital ulcers status in the past year* |  |  |  |  |  |  |
| No | 30.6 (649/2120) | 9.6 (1363/14 174) | 3.18 (2.93-3.45) | <0.001 | 1.45 (1.32-1.59) | <0.001 |
| Yes | 46.7 (140/300) | 22.0 (186/844) | 2.12 91.78-2.52) | <0.001 | 1.18 (0.97-1.43) | 0.105 |
| **Number of lifetime sexual partner(s)** |  |  |  |  |  |  |
| Not sexually active | - | 0.8 (26/3230) | - | - | - | - |
| 1-2 | 6.8 (3/44) | 3.1 (77/2479) | 2.19 (0.72-6.69) | 0.167 | 1.43 (0.46-4.50) | 0.536 |
| 3-4 | 36.4 (561/1542) | 22.0 (678/3089) | 1.66 (1.51-1.82) | <0.001 | 1.22 (0.10-1.35) | <0.001 |
| 5-10 | 19.1 (29/152) | 8.1 (224/2751) | 2.34 (1.65-3.33) | <0.001 | 1.25 (0.86-1.82) | 0.234 |
| 11-20 | 25.2 (100/397) | 14.4 (415/2879) | 1.75 (1.44-2.12) | <0.001 | 1.23 (1.01-1.51) | 0.039 |
| >20 | 33.5 (56/167) | 21.1 (93/440) | 1.59 (1.20-2.10) | 0.001 | 1.41 (1.03-1.92) | 0.032 |
| a lot/many (>3) <sup>§</sup> | 33.3 (39/117) | 24.3 (33/136) | 1.37 (0.93-2.04) | 0.113 | 1.11 (0.75-1.65) | 0.595 |
| **Number of sexual partner(s) in the past year** |  |  |  |  |  |  |
| Not sexually active | - | 3.0 (129/4342) | - | - | - | - |
| 1 | 38.8 (114/294) | 13.1 (794/6072) | 2.97 (2.53-3.47) | <0.001 | 1.46 (1.21-1.75) | <0.001 |
| 2 | 35.3 (235/666) | 14.1 (437/3095) | 2.50 (2.18-2.86) | <0.001 | 1.44 (1.24-1.68) | <0.001 |
| 3-5 | 31.0 (309/998) | 12.5 (169/1343) | 2.46 (2.08-2.91) | <0.001 | 1.47 (1.22-1.77) | <0.001 |
| >5 | 26.9 (42/156) | 12.8 (5/39) | 2.10 (0.89-4.97) | 0.091 | 1.68 (0.73-3.85) | <0.001 |
| a lot/many (>3) <sup>§</sup> | 29.1 (89/306) | 11.8 (15/127) | 2.46 (1.48-4.09) | <0.001 | 1.38 (0.82-2.31) | 0.226 |

Abbreviations: FBSW, female bar or sex worker; PR, prevalence ratio;
\*Genital ulcers in the past year for round 16-18, and genital ulcers in the past month for round 19
<sup>§</sup>An option in the questionnaire for individuals with more than three sexual partners but cannot recall the exact number as a separate category
Note: The adjusted model included adjustments for age in years, community type, migration status, education level, socioeconomic status, and marital status among participants with complete information on covariates (N<sub>individual</sub> = 17 349)

HIV seroprevalence was higher among men with FBSW partners compared to those not sexually active or sexually active without FBSWs, with the highest seroprevalence in those with FBW partners only(**Supplementary_Figures_2-3**). Among study visits where men reported four sexual partners in the past year(n=2,601), HIV seroprevalence increased by the number of FBSWs reported, reaching 37.3% at visits in which all four partners were FBSWs (*P*_trend_<0.001) (**Figure_1B**). Among partnerships involving FBSWs (n=5,505), HIV seroprevalence increased by the length of partnership (*P*_trend_<0.001)(**Figure_1C**).

### HIV incidence

Among 12,695 HIV-seronegative male participants at their first visit, 8,078 (63.6%) had at least one follow-up visit. Characteristics of included and excluded visits are detailed in **Supplementary_Table_5**, with similar proportions of excluded visits for men with (32.9%) and without (33.4%) FBSW partners, though differences existed by sociodemographic and behavioral factors.

Over 27,396 person-years of follow-up, 154 HIV seroconversions occurred, with an overall incidence of 0.56/100 person-years: 0.44/100 among men without FBSW partners and 1.93/100 among those with FBSW partners (IRR=4.39,95%CI=3.04-6.16; age-adjusted IRR=4.46,95% CI=3.09-6.32)(**Table_4**). The age-adjusted IRR was similar with censoring weights (IRR=4.40,95%CI=3.04-6.24) and when restricted to sexually active men (IRR=3.69, 95%CI=2.54-5.26). Stratified by community type, associations were stronger in inland than fishing communities (IRR=3.66vs.1.95,*P*_interaction_=0.030). From 2013 to 2020, HIV incidence declined from 0.49 to 0.34/100 person-years among men without FBSW partners and from 2.04 to 1.75/100 among those with FBSW partners, yet disparities widened, with age-adjusted IRR rising from 4.06 (2013–2016) to 6.19 (2016–2020).

**Table 4.**
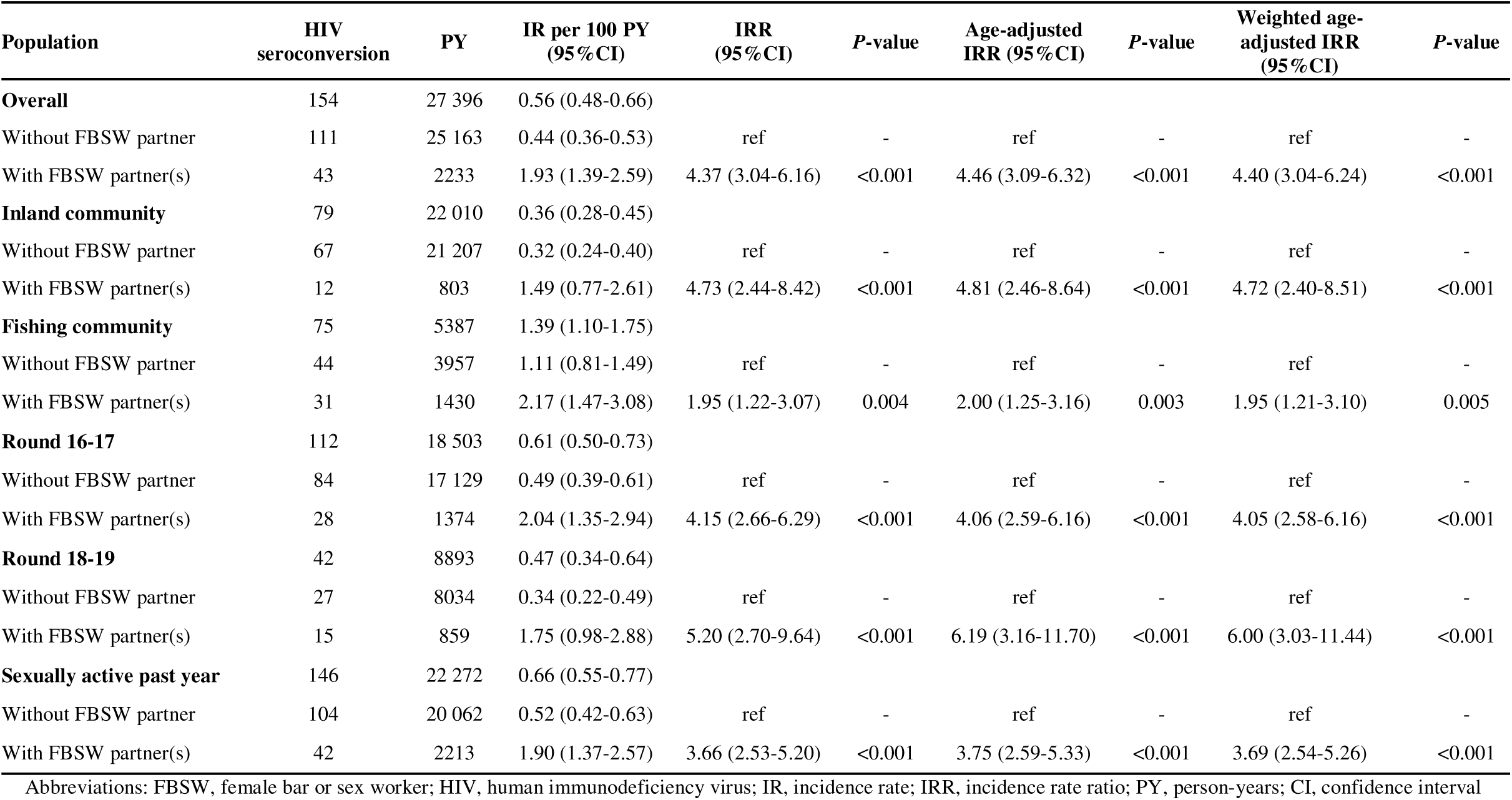
Incidence rates and ratios of HIV seroconversion by self-reported engagement with female bar or sex worker (FBSW) partners <u>at either the start or end of a visit interval</u> among male participants in the Rakai Community Cohort Study, Uganda, 2013-2020 (N_individual_ = 8078)

In sensitivity analyses comparing male participants who ever reported FBSW to those who never reported FBSW partners, HIV incidence differences were similar(**Supplementary_Table_6**).

### HIV viral suppression

During the study period, 2,468 men tested HIV-seropositive, with viral load data at 5,195/5,214 (99.6%) visits. Overall, viral suppression increased over time but was slightly lower at visits reporting FBSW partners compared to visits without (70.2%vs.73.4%;PR=0.95,95% CI=0.91-1.00), a difference primarily observed in fishing communities (PR=0.93,95% CI=0.88-0.98)(**Supplementary_Table_7, Supplementary_Figure_4**).

### Population prevalence of HIV viremia

**Figure 2** shows the population prevalence and prevalence ratios of HIV viremia by survey round and FBSW partner exposure. Between surveys 16 and 19, viremia prevalence declined from 18.8% to 3.8% among men with FBSW partners and from 5.2% to 1.9% among those without, though disparities persisted, with an overall PR of 1.61 (95%CI=1.41-1.84) across the study period, accounting for repeated measures and adjusted for age and community type. While fishing communities consistently showed a higher population prevalence of HIV viremia than inland communities, the disparity between men with and without FBSW partners was more pronounced in inland communities.

### HIV awareness, PrEP use, and condom use

Among 1,201 men living with HIV in survey round 19 (2018–2020), HIV serostatus awareness was slightly higher among those with FBSW partners (95.3%) than those without (91.3%)(*P*=0.029) (**Supplementary_Table_8**). Of 6,910 HIV-seronegative men who self-reported PrEP use status in survey round 19, 73.9% (465/629) with FBSW partners had heard of PrEP, compared to 69.2% (4,345/6,281) among those without(*P*=0.014). PrEP use was also higher among men with FBSW partners, with 10.6% reporting ever using it and 5.7% reporting current use, compared to 2.0% and 1.0%, respectively, for men without(*P*<0.001)(**Supplementary_Table_9**). Regarding condoms, HIV seroprevalence was significantly lower in FBSW partnerships with consistent condom use compared to those with never use (31.8%vs.40.1%, *P*< 0.001). Relatedly, males with HIV and FBSW partners were less likely to consistently use condoms in their partnerships compared to HIV-seronegative males (43.1%vs.51.6%, *P*<0.001)(**Supplementary_Table_10**).

## Discussion

In this population-based longitudinal study in Uganda, 13.9% of men self-reported having FBSW partners. Men with FBSW partners tended to be older, have lower SES, be more likely to be previously married, and reside in Lake Victoria fishing communities. They also exhibited greater levels of HIV-associated risk behaviors, including having more sexual partners and alcohol use during sex. Men with FBSW partners had a significantly elevated HIV seroprevalence and faced a markedly higher risk of HIV acquisition compared to the local male population without FBSW partners. Despite comparable levels of viral suppression among men with HIV, those with FBSW partners had a substantially higher population prevalence of HIV viremia. Encouragingly, men with FBSW partners exhibited a greater awareness of HIV serostatus and familiarity with PrEP. However, PrEP uptake was alarmingly low, with only ∼6% reporting current use, despite many being eligible due to exchanging money for sex or having multiple partners of unknown HIV status.^22^ Taken together, these data suggest that men who have sex with FBSWs remain at substantial risk of HIV acquisition and onward transmission. Tailored interventions coupled with community-engaged approaches to improve PrEP delivery and use are urgently needed to reduce the burden of HIV in this population.

Our study estimated an HIV seroprevalence of ∼33% among Ugandan males with FBSW partners, which was three times higher than that of the general male population. While men with FBSW partners exhibited higher levels of HIV-associated risk factors compared to those without, seroprevalence increased with greater numbers of FBSW partners reported and with increasing duration of partnership with an FBSW partner. Notably, differentials in HIV seroprevalence between those with and without FBSW partners were greater in lower-prevalence inland communities, which may be due to saturation effects in extremely high-burden fishing communities. The three-fold higher prevalence of HIV among male partners of FBSW in this study is substantially greater than the 1.62 pooled prevalence ratio reported in the 2022 meta-analysis of East African men who had ever paid for sex.^17^ Differences may be a result of our study directly measuring sex with FBSW partners rather than a combination of these and less risky transactional sex relationships. Notably, a study of 162 regular male partners of FSWs in Kampala, Uganda reported an HIV prevalence of 40%, similar to our findings.^13^

We also identified a significant disparity in HIV incidence between men with and without FBSW partners. While a higher proportion of men with FBSW partners were aware of their HIV serostatus (95.3%) and had heard of PrEP (73.9%), PrEP uptake was low, with fewer than 6% reporting current use. Additionally, nearly half of male partnerships with FBSWs lacked mutual awareness of partners’ HIV serostatus, a factor shown to hinder effective risk reduction and HIV prevention efforts.^23^ A similar proportion of partnerships with FBSWs included no or inconsistent condom use. These gaps between awareness and the use of prevention tools highlight a critical need for tailored strategies. On-demand or long-acting injectable PrEP modalities could significantly reduce risk but would require effective engagement with these men to improve uptake and ensure persistent use.^24–26^

Although viral suppression among men living with HIV improved for those with and without FBSW partners over the study period, men with FBSW partners exhibited a higher population prevalence of HIV viremia due to their high background seroprevalence. These findings underscore their potential role in ongoing virus transmission, particularly given that 77% of study visits involving sex with at least one FBSW also included non-FBSWs. We have previously reported a growing gender disparity in HIV acquisition and transmission risks in this same Ugandan setting, with an increasing proportion of transmission attributed to men as overall HIV incidence declines.^27^ Modeling studies suggest that partners of members of key populations may account for up to a quarter of HIV infections and that scaling up intervention coverage among these subgroups could substantially interrupt transmission.^28–30^ Notably, partnerships with FBWs tended to be longer-term and were more often identified as girlfriend or committed relationships, compared to those with FSWs, which tended to be one-time partnerships. While interventions that focus on venues may be effective for HIV prevention and care among women regardless of whether they identify as sex workers or bar workers, tailored approaches for males in long-term relationships with FBWs may help increase the use of prevention methods.

This study has several limitations. First, only the four most recent sexual partnerships and behaviors were self-reported, which may be subject to recall and social desirability biases, potentially leading to underreporting of FBSW partners and high-risk behaviors, particularly given the stigma surrounding transactional sex and HIV status disclosure in this region.^31–33^ Second, while our study included a large and diverse sample across multiple survey rounds, the findings may not be generalizable to all regions of Africa or other populations with different contexts. Third, we cannot entirely rule out selection bias due to non-participation; however, no significant differences in loss to follow-up were observed between those with and without FBSW partners, and analyses incorporating censoring weights did not change study findings. Lastly, the most recent metrics were reported in 2019-20 and may not fully reflect current epidemic trends or patterns of service use.

In conclusion, our findings highlight an urgent need for prioritizing HIV prevention and care services for men with FBSW partners in Uganda. Expanding PrEP access, improving condom distribution, enhancing risk reduction counseling, and promoting regular HIV testing and status disclosure within this population could avert new infections in this setting. Prioritizing these men in HIV prevention and control strategies could play a pivotal role in reducing HIV incidence among them and their sexual partners while achieving epidemic control.

## Data Availability

All data produced in the present study are available upon reasonable request to the Rakai Health Sciences Program

## Contributors

XF, KG, AART, and RS conceived and designed the study. FN, GK, GN, AN, HN, RMG, DS, and VS collected and managed the data. XF and KG conducted statistical methods and analyses. XF, KG, LWC, AW, CEK, and EUP contributed to the interpretation of the findings. XF, KG, and AART drafted the manuscript, and all authors participated in revising it. All authors had access to all of the data in the study and read and approved the final manuscript for publication.

## Data sharing

De-identified Rakai Community Cohort Study data can be provided to interested parties subject to the completion of the Rakai Health Sciences Program data request form and the signing of a Data Transfer Agreement. Inquiries should be directed to.

## Acknowledgment

We sincerely thank all RCCS participants for their time and valuable contributions to this study. We also acknowledge the dedication and hard work of the data collection, laboratory, service linkage, and data management staff at RHSP. We thank members of the Johns Hopkins Bloomberg School of Public Health and NIAID Laboratory of Immunoregulation International HIV/STD Section at the Johns Hopkins University School of Medicine for their helpful contributions to this work. This study was supported in part by the National Institutes of Health (NIH) National Institute of Allergy and Infectious Diseases (NIAID) (U01AI075115, R01AI087409, U01AI100031, R01AI110324, R01AI114438, K25AI114461, R01AI123002, R01AI128779, R01AI143333, R21AI145682, R01AI155080, ZIAAI001040), NIH National Institute of Child Health and Development (R01HD050180, R01HD070769, R01HD091003), NIH National Heart, Lung, and Blood Institute (R01HL152813), National Institute of Diabetes, Digestive, and Kidney Diseases (R01DK131926), the Fogarty International Center (D43TW009578, D43TW010557), the Johns Hopkins University Center for AIDS Research (P30AI094189). This project was supported by the National Institutes of Health and in part by the President’s Emergency Plan for AIDS Relief (PEPFAR) through the U.S. Centers for Disease Control and Prevention (U.S. CDC) cooperative agreement numbers NU2GGH002009 and NU2GGH000817. This project was also supported in part by the Division of Intramural Research, NIAID, NIH.

