## Supplementary Materials for "HIV seroprevalence, incidence, and viral suppression among Ugandan males with bar or sex worker partners: a population-based study"

### **Supplementary Statistical Methods**

All statistical analyses were conducted using the Stata 15.1 and R statistical software (version 4.2.1).

#### Calculation on inverse probability of censoring weight (IPCW)

For each participant, logistic regression was used to estimate the probability of follow-up, considering factors including age, community type, migration status, education level, SES, marital status, and number of lifetime and past-year sexual partners. We assigned each individual a weight of 1/probability and stabilized these weights, ensuring a mean weight of 1 in our study population. The IPCW was then incorporated into the Poisson regressions to calculate incidence rate ratios of HIV infection between FBWs and non-FBWs, accounting for selection bias due to differences between participants observed in the incidence cohort and those lost to follow-up.

### **Supplementary Tables and Figures**

#### Supplementary Table 1. Participation rates by survey round of males in the Rakai Community Cohort Study, Uganda, 2013-2020 (N _individual_ = 17 438)

| **Survey round** | **Number of participants** | **Number of eligible men** | **Participation rate (%)** |
| --- | --- | --- | --- |
| 16 | 8308 | 13 847 | 60.0 |
| 17 | 8866 | 14 758 | 60.1 |
| 18 | 9243 | 14 977 | 61.7 |
| 19 | 8856 | 15 380 | 57.6 |
| Overall | 17 438 | 26 871 | 64.9 |

Note: participation rates were calculated as the proportion of participants in our study among males eligible in the community at the time of the survey

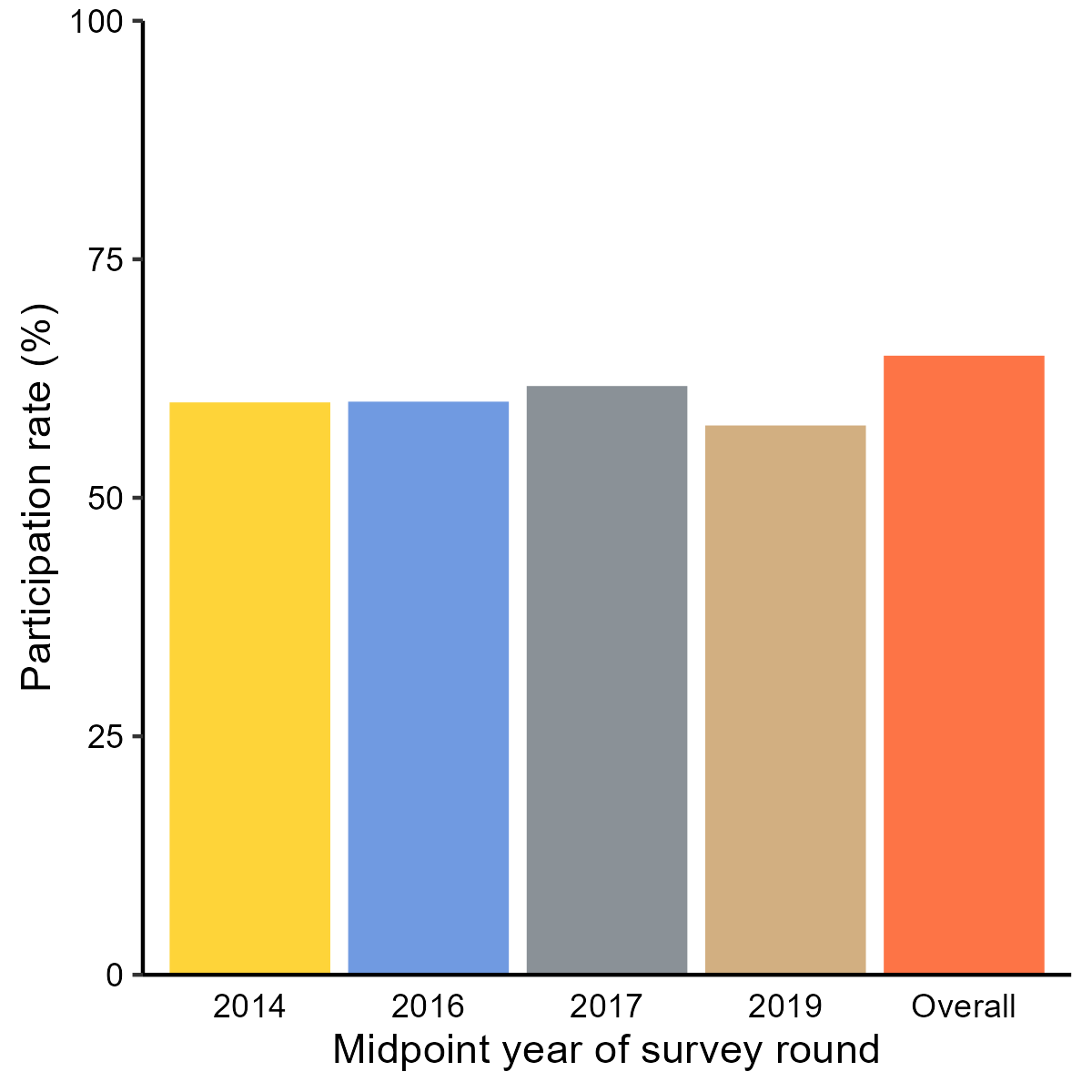

#### Supplementary Figure 1. Participation rates by survey round of the Rakai Community Cohort Study, Uganda, 2013-2020 (N _individual_ = 17 438)

#### Supplementary Table 2. The proportion of male participants reporting engagement with female bar or sex worker (FBSW) partners past year by community type and survey round in the Rakai Community Cohort Study, Uganda, 2013-2020 (N _individual_ = 17 438)

| **Community type** | **Midpoint year of survey round** | **Total number** | **Not sexually active past year** | **Sexually active past year** | | | |
| --- | --- | --- | --- | --- | --- | --- | --- |
|  |  |  |  | **Without FBSW partner** | **With FBSW partner(s)** | **With FBW partner(s)** | **With FSW partner(s)** |
| Inland | 2014 | 6247 | 1453 (23.3%) | 4598 (73.6%) | 196 (3.1%) | 192 (3.1%) | 20 (0.3%) |
| Inland | 2016 | 6705 | 1284 (19.1%) | 5196 (77.5%) | 225 (3.4%) | 202 (3.0%) | 51 (0.8%) |
| Inland | 2017 | 6711 | 1345 (20.0%) | 5089 (75.8%) | 277 (4.1%) | 221 (3.3%) | 94 (1.4%) |
| Inland | 2019 | 6745 | 1355 (20.1%) | 5057 (75.0%) | 333 (4.9%) | 295 (4.4%) | 91 (1.3%) |
| Fishing | 2014 | 2061 | 138 (6.7%) | 1414 (68.6%) | 509 (24.7%) | 388 (18.8%) | 222 (10.8%) |
| Fishing | 2016 | 2161 | 129 (6.0%) | 1448 (67.0%) | 584 (27.0%) | 448 (20.7%) | 312 (14.4%) |
| Fishing | 2017 | 2532 | 148 (5.8%) | 1642 (64.8%) | 742 (29.3%) | 544 (21.5%) | 415 (16.4%) |
| Fishing | 2019 | 2111 | 106 (5.0%) | 1404 (66.5%) | 601 (28.5%) | 411 (19.5%) | 333 (15.8%) |

Abbreviations: FBSW, female bar or sex worker

#### Supplementary Table 3. Characteristics of sexual partnership by involvement with female bar or sex workers among males with FBSW partners in the Rakai Community Cohort Study, Uganda, 2013-2020 (N _individual_ = 2420)

| **Partnership characteristics** | **Partnership with FBW(s)** | **Partnership with FSW(s)** | ***P-*value** |
| --- | --- | --- | --- |
|  | **N _partnership_ = 3408** | **N _partnership_ = 2766** |  |
| **Length of relationship in days, median (IQR)** | 30.0 (0.0, 275.0) | 0.0 (0.0, 0.0) | <0.001 |
| **Relationship type** |  |  | <0.001 |
| Current wife | 81 (2.4%) | 2 (0.1%) |  |
| Current consensual partner | 487 (14.3%) | 2 (0.1%) |  |
| Girlfriend | 1446 (42.4%) | 107 (3.9%) |  |
| Occasional or casual friend | 550 (16.1%) | 249 (9.0%) |  |
| Client/sex worker | 814 (23.9%) | 2385 (86.3%) |  |
| Others | 30 (0.9%) | 20 (0.7%) |  |
| **Relationship ongoing** |  |  | <0.001 |
| Yes | 1160 (34.0%) | 80 (2.9%) |  |
| No | 2240 (65.7%) | 2678 (96.9%) |  |
| Do not know | 8 (0.2%) | 7 (0.3%) |  |
| **Condom use** |  |  | <0.001 |
| Never | 1716 (50.4%) | 752 (27.2%) |  |
| Inconsistent | 517 (15.2%) | 108 (3.9%) |  |
| Always | 1173 (34.4%) | 1901 (68.8%) |  |
| Do not know | 2 (0.1%) | 4 (0.1%) |  |
| **Partner's HIV status was ever known by male participant** | |  | <0.001 |
| Yes | 1070 (31.4%) | 171 (6.2%) |  |
| No | 2338 (68.6%) | 2594 (93.8%) |  |
| Do not know | 0 (0.0%) | 0 (0.0%) |  |
| **Male participant informed partner of his HIV status** | |  | <0.001 |
| Yes | 1030 (30.2%) | 200 (7.2%) |  |
| No | 1973 (57.9%) | 2418 (87.5%) |  |
| Never tested/got HIV results | 186 (5.5%) | 145 (5.2%) |  |
| Received couple counseling | 219 (6.4%) | 2 (0.1%) |  |
| Do not remember | 0 (0.0%) | 0 (0.0%) |  |
| **Male participant uses alcohol before sex with partner*** | **N _partnership_ = 2516** | **N _partnership_ = 1996** | <0.001 |
| Yes | 1693 (67.3%) | 1071 (53.7%) |  |
| No | 823 (32.7%) | 925 (46.3%) |  |
| **Partner uses alcohol before sex act with male participant*** | **N _partnership_ = 2516** | **N _partnership_ = 1996** | <0.001 |
| Yes | 1669 (66.3%) | 882 (44.2%) |  |
| No | 847 (33.7%) | 1114 (55.8%) |  |
| Do not know | 0 (0.0%) | 0 (0.0%) |  |

*Among male participants in the Rakai Community Cohort Study, Uganda, 2013-2018 (N _individual_ = 1900)

Abbreviations: FBSW, female bar or sex worker; IQR, interquartile range; RCCS, Rakai Community Cohort Study; HIV, human immunodeficiency virus

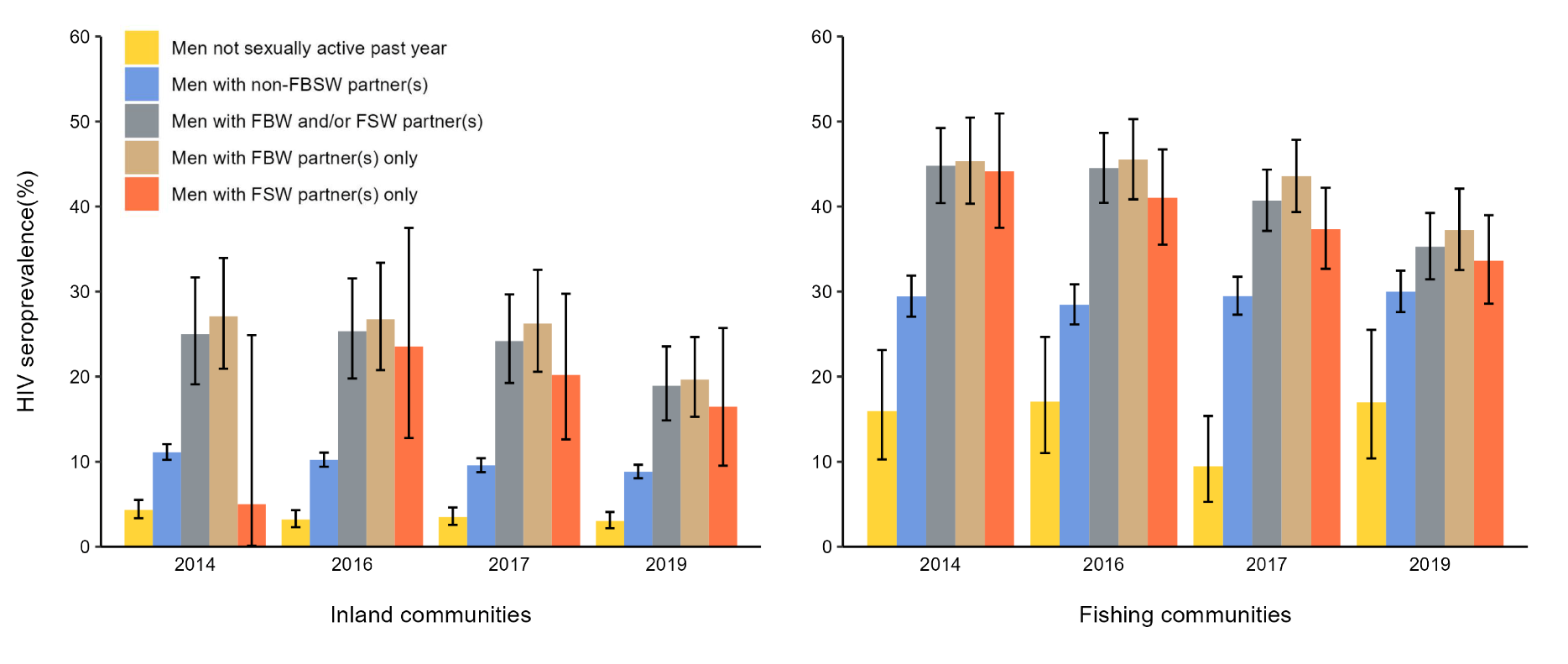

#### Supplementary Figure 2. HIV seroprevalence at each survey round by self-reported engagement with female bar or sex worker (FBSW) partners among male participants in the Rakai Community Cohort Study, Uganda, 2013-2020 (N _individual_ = 17 438)

#### Supplementary Table 4. HIV seroprevalences and prevalence ratios at baseline by self-reported engagement with female bar or sex worker (FBSW) partners among male participants in the Rakai Community Cohort Study, Uganda, 2013-2020 (N _individual_ = 17 438)

| **Population** | **Seroprevalence**  **(case/total)** | **Model 1** | | **Model 2** | | **Model 3** | | **Model 4** | |
| --- | --- | --- | --- | --- | --- | --- | --- | --- | --- |
|  |  | **PR**  **(95%CI)** | ***P-*value** | **Adjusted PR**  **(95%CI)** | ***P-*value** | **Adjusted PR**  **(95%CI)** | ***P-*value** | **Adjusted PR**  **(95%CI)** | ***P-*value** |
| **Overall** |  |  |  |  |  |  |  |  |  |
| Without FBSW partner | 10.3% (1549/15 018) | ref |  | ref |  | ref |  | ref |  |
| With FBSW partner(s) | 32.6% (789/2420) | 3.16 (2.93-3.40) | <0.001 | 2.46 (2.28-2.65) | <0.001 | 1.44 (1.32-1.57) | <0.001 | 1.38 (1.24-1.52) | <0.001 |
| **Inland community** |  |  |  |  |  |  |  |  |  |
| Without FBSW partner | 7.5% (925/12 309) | ref |  | ref |  | ref |  | ref |  |
| With FBSW partner(s) | 21.0% (172/820) | 2.79 (2.41-3.23) | <0.001 | 1.89 (1.63-2.19) | <0.001 | 1.58 (1.35-1.84) | <0.001 | 1.48 (1.24-1.76) | <0.001 |
| **Fishing community** |  |  |  |  |  |  |  |  |  |
| Without FBSW partner | 23.0% (624/2709) | ref |  | ref |  | ref |  | ref |  |
| With FBSW partner(s) | 38.6% (617/1600) | 1.67 (1.53-1.84) | <0.001 | 1.57 (1.43-1.72) | <0.001 | 1.40 (1.26-1.52) | <0.001 | 1.29 (1.14-1.44) | <0.001 |

Abbreviation: FBSW, female bar or sex worker; PR, prevalence ratio

**Model 1** provided unadjusted results; **Model 2** adjusted for age in years; **Model 3** additionally adjusted for (community type), migration status, education attainment, socioeconomic status, and marital status among participants with complete data on covariates (N _individual_ = 17 349); **Model 4** additionally adjusted for the number of lifetime sexual partners at the time of visit, circumcision status, and the past-year self-reported number of sexual partners, genital ulcer (GUD) status, transactional sex, non-marital partnerships, and consistent condom usage with non-marital partners among participants from survey rounds 15-19 who were sexually active in the past year and had complete data on covariates (N _individual_ = 13 562).

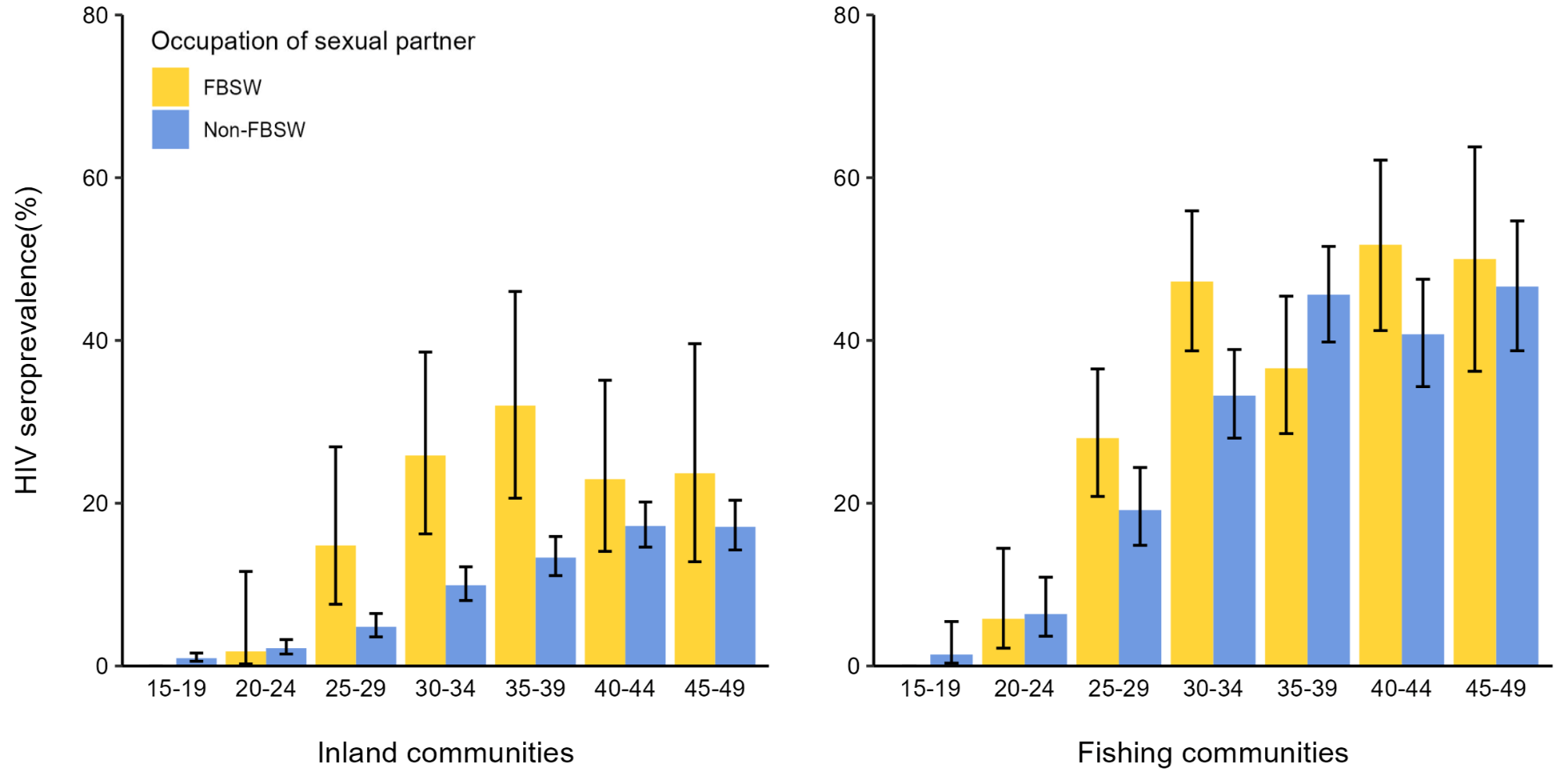

Abbreviations: FBSW, female bar or sex worker; HIV, human immunodeficiency virus

#### Supplementary Figure 3. HIV seroprevalence by self-reported engagement with female bar or sex worker (FBSW) partners in the inland and fishing communities among male participants of the survey round 19 of Rakai Community Cohort Study, Uganda, 2018-2020 (N _individual_ =10 414)

#### Supplementary Table 5. Characteristics of individual visits included in the incidence cohort and excluded due to loss to follow-up among male participants in the Rakai Community Cohort Study, Uganda, 2013-2020 (N _visit_ = 22 404)

| **Characteristics** | **Included visits** | **Excluded visits** | ***P-*value** | |
| --- | --- | --- | --- | --- |
|  | **N _visit_ = 14 932** | **N _visit_ = 7472** |  |  |
| **Bar or sex work** |  |  | 0.690 | |
| No | 13 680 (66.6%) | 6857 (33.4%) |  | |
| Yes | 1252 (67.1%) | 615 (32.9%) |  | |
| **Age category** |  |  | <0.001 | |
| 15-19 | 3013 (61.1%) | 1918 (38.9%) |  | |
| 20-24 | 2528 (60.5%) | 1648 (39.5%) |  | |
| 25-29 | 2479 (67.0%) | 1221 (33.0%) |  | |
| 30-34 | 2260 (72.2%) | 872 (27.8%) |  | |
| 35-39 | 2072 (75.0%) | 684 (24.8%) |  | |
| 40-44 | 1742 (79.6%) | 447 (20.4%) |  | |
| 44-49 | 838 (55.1%) | 682 (44.9%) |  | |
| **Community type** |  |  | 0.170 | |
| Inland | 11 909 (66.9%) | 5901 (33.1%) |  | |
| Fishing | 3023 (65.8%) | 1571 (34.2%) |  | |
| **Migration** |  |  | <0.001 | |
| Long-term resident | 13 035 (70.6%) | 5433 (29.4%) |  | |
| Migrants | 1897 (48.2%) | 2039 (51.8%) |  | |
| **Education attainment** |  |  | <0.001 | |
| None | 396 (64.1%) | 222 (35.9%) |  | |
| Primary | 9754 (64.1%) | 4486 (31.5%) |  | |
| Secondary | 3811 (68.5%) | 2200 (31.5%) |  | |
| Technical/University | 970 (63.4%) | 563 (36.6%) |  | |
| Missing | 1 (50%) | 1 (50%) |  | |
| **Socioeconomic status** | |  | <0.001 | |
| Lowest | 2201 (67.6%) | 1053 (32.4%) |  | |
| Low-middle | 2254 (70.3%) | 951 (29.7%) |  | |
| High-middle | 4196 (66.2%) | 2140 (33.8%) |  | |
| Highest | 6231 (65.4%) | 3294 (34.6%) |  | |
| Missing | 50 (59.5%) | 34 (40.5%) |  | |
| **Marital status** |  |  | <0.001 | |
| Currently married | 8002 (70.7%) | 3323 (29.3%) |  | |
| Previously married | 1492 (68.4%) | 3459 (31.6%) |  | |
| Never married | 5438 (61.1%) | 3459 (38.9%) |  | |
| **Number of lifetime sexual partners** | |  | <0.001 | |
| 0 | 2098 (62.7%) | 1250 (37.3%) |  | |
| 1-2 | 2405 (64.5%) | 1323 (35.5%) |  | |
| 3-4 | 3718 (67.1%) | 1825 (32.9%) |  | |
| 5-10 | 2845 (67.6%) | 1362 (32.4%) |  | |
| 11-20 | 3156 (69.2%) | 1403 (30.8%) |  | |
| >20 | 515 (71.5%) | 205 (28.5%) |  | |
| a lot/many (>3) ^§^ | 186 (64.8%) | 101 (35.2%) |  | |
| Missing | 9 (75.0%) | 3 (25%) |  | |
| **Number of sexual partners in the past year** | | | | <0.001 |
| 0 | 2987 (62.0%) | 1828 (38.0%) |  | |
| 1 | 6228 (68.1%) | 2924 (32.0%) |  | |
| 2 | 3432 (67.9%) | 1620 (32.1%) |  | |
| 3-5 | 1707 (66.2%) | 871 (33.8%) |  | |
| >5 | 83 (63.9%) | 47 (36.2%) |  | |
| a lot/many (>3) ^§^ | 266 (65.2%) | 142 (34.8) |  | |
| Missing | 229 (85.1%) | 40 (14.9%) |  | |

^§^ An option in the questionnaire for individuals with more than three sexual partners but cannot recall the exact number as a separate category

#### Supplementary Table 6. Incidence rates and incidence rate ratios of HIV seroconversion by self-reported ever engagement with female bar or sex worker (FBSW) partners among male participants in the Rakai Community Cohort Study, Uganda, 2013-2020 (N _individual_ = 8078)

| **Population** | **HIV seroconversion** | **Person-years** | **IR per 100 PY (95%CI)** | **IRR**  **(95%CI)** | ***P-*value** | **Age-adjusted IRR**  **(95%CI)** | ***P-*value** |
| --- | --- | --- | --- | --- | --- | --- | --- |
| **Overall** | 154 | 27 396 | 0.56 (0.48-0.66) |  |  |  |  |
| Without FBSW partner | 92 | 23 422 | 0.39 (0.32-0.48) | ref | - | ref | - |
| With FBSW partner(s) | 62 | 3975 | 1.56 (1.20-2.00) | 4.37 (3.04-6.16) | <0.001 | 4.46 (3.09-6.32) | <0.001 |
| **Inland community** | 79 | 22 010 | 0.36 (0.28-0.45) |  |  |  |  |
| Without FBSW partner | 62 | 20 318 | 0.31 (0.23-0.39) | ref | - | ref | - |
| With FBSW partner(s) | 17 | 1692 | 1.00 (0.59-1.61) | 4.73 (2.44-8.42) | <0.001 | 4.81 (2.46-8.64) | <0.001 |
| **Fishing community** | 75 | 5387 | 1.39 (1.10-1.75) |  |  |  |  |
| Without FBSW partner | 30 | 3104 | 0.97 (0.65-1.38) | ref | - | ref | - |
| With FBSW partner(s) | 45 | 2283 | 1.97 (1.44-2.64) | 1.95 (1.22-3.07) | 0.004 | 2.00 (1.25-3.16) | 0.003 |

Abbreviations: FBSW, female bar or sex worker; HIV, human immunodeficiency virus; IR, incidence rate; CI, confidence interval; IRR, incidence rate ratio; PY, person-years

#### Supplementary Table 7. Prevalence and prevalence ratios of HIV viral suppression by self-reported engagement with female bar or sex worker (FBSW) partners among HIV seropositive visits of male participants in the Rakai Community Cohort Study, Uganda, 2013-2020 (N _visit_ = 5195)

| **Population** | **Viral suppression, %**  **(case/total)** | **PR (95%CI)** | ***P*-value** | **Age-adjusted PR (95%CI)** | ***P*-value** |
| --- | --- | --- | --- | --- | --- |
| **Overall** | 72.7 (3774/5195) |  |  |  |  |
| Without FBSW partner | 73.4 (2904/3955) | ref | - | ref | - |
| With FBSW partner(s) | 70.2 (870/1240) | 0.95 (0.91-1.00) | 0.036 | 0.98 (0.94-1.02) | 0.270 |
| **Inland community** | 73.2 (1748/2389) |  |  |  |  |
| Without FBSW partner | 72.9 (1568/2150) | ref | - | ref | - |
| With FBSW partner(s) | 83.0 (180/239) | 1.03 (0.95-1.12) | 0.500 | 1.04 (0.95-1.12) | 0.403 |
| **Fishing community** | 72.2 (2026/2806) |  |  |  |  |
| Without FBSW partner | 74.0 (1336/1805) | ref | - | ref | - |
| With FBSW partner(s) | 68.9 (690/1001) | 0.93 (0.88-0.98) | 0.004 | 0.95 (0.90-0.99) | 0.031 |

Abbreviations: FBSW, female bar or sex worker; PR, prevalence ratio; CI, confidence interval.

Note: Prevalence ratios were calculated by Poisson regression models with generalized estimating equations (GEE), using an exchangeable correlation structure and robust standard errors to account for within-subject correlations over time

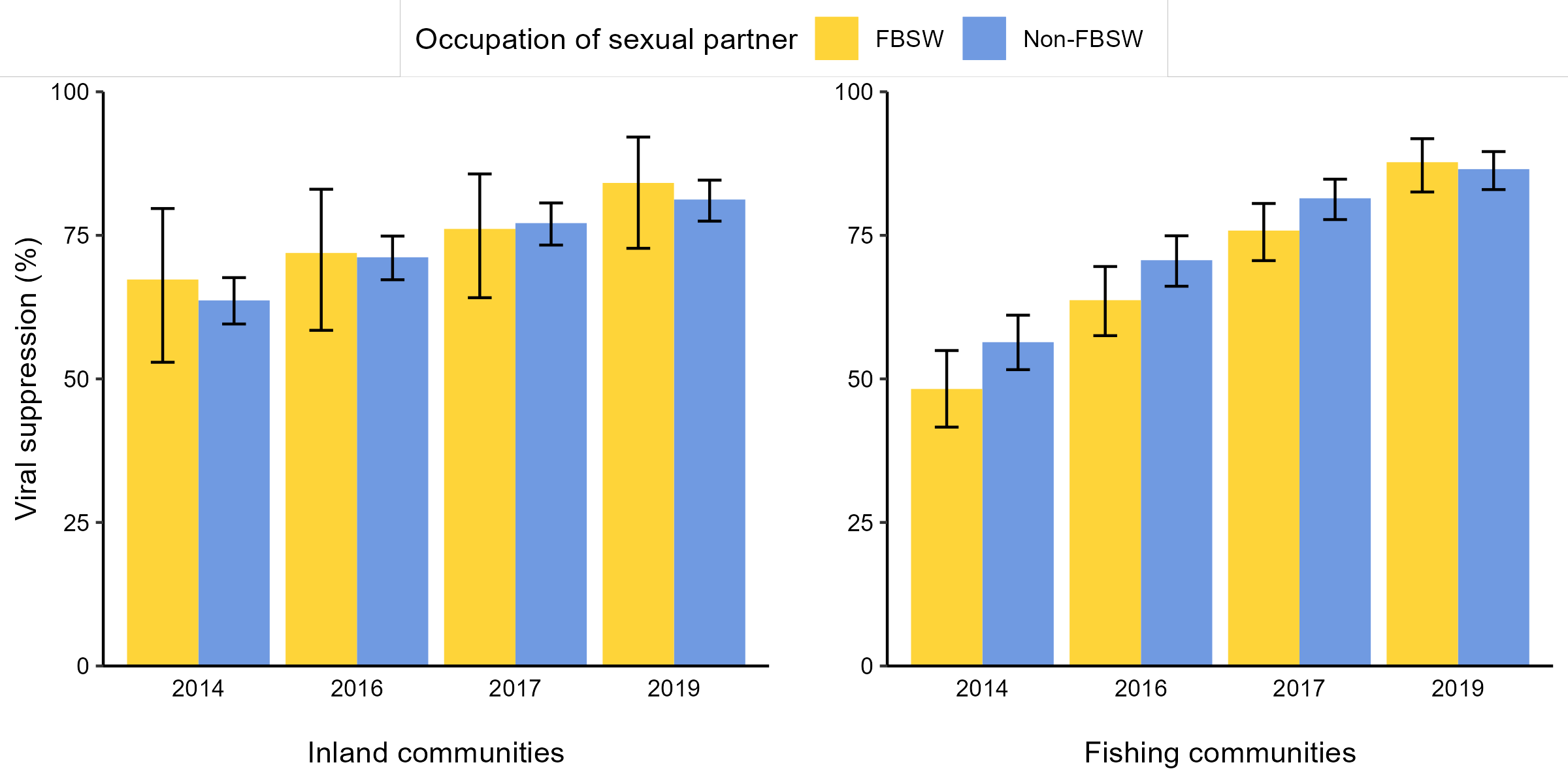

#### **Supplementary Figure 4**. HIV viral suppression at each survey round by self-reported engagement with female bar or sex worker (FBSW) partners among HIV seropositive visits of male participants in the Rakai Community Cohort Study, Uganda, 2013-2020 (N _visit_ = 5195)

#### Supplementary Table 8. Awareness of HIV serostatus among male participants living with HIV in survey round 19 of the Rakai Community Cohort Study, Uganda, 2018-2020 (N _individual_ = 1201)

| **Awareness of HIV serostatus** | **Without FBSW partners** | **With FBSW partners** | ***P-*value** |
| --- | --- | --- | --- |
|  | **N _individual_ = 926** | **N _individual_ = 275** |  |
| No | 81 (8.7%) | 13 (4.7%) | 0.029 |
| Yes | 845 (91.3%) | 262 (95.3%) |  |

Abbreviations: FBSW, female bar or sex worker; HIV, human immunodeficiency virus

#### Supplementary Table 9. PrEP awareness and use among HIV seronegative male participants in survey round 19 of the Rakai Community Cohort Study, Uganda, 2018-2020 (N _individual_ = 6910)

| **Characteristics** | **Without FBSW partners** | **With FBSW partners** | ***P-*value** |
| --- | --- | --- | --- |
|  | **N _individual_ = 6281** | **N _individual_ = 629** |  |
| **PrEP awareness** |  |  | 0.014 |
| No | 1936 (30.8%) | 164 (26.1%) |  |
| Yes | 4345 (69.2%) | 465 (73.9%) |  |
| **PrEP use** |  |  | <0.001 |
| Never | 6153 (98.0%) | 562 (89.4%) |  |
| Previous | 65 (1.0%) | 31 (4.9%) |  |
| Current | 63 (1.0%) | 36 (5.7%) |  |

Abbreviations: FBSW, female bar or sex worker; HIV, human immunodeficiency virus; PrEP, Pre-exposure prophylaxis

#### Supplementary Table 10. Prevalence of condom use among partnerships involving and not involving female bar or sex workers (FBSWs), stratified by HIV serostatus among male participants in the Rakai Community Cohort Study, Uganda, 2013-2020 (N _individual_ = 14 740)

| **Partnership** | **HIV serostatus** | **Condom use** | | | **Total** |
| --- | --- | --- | --- | --- | --- |
|  |  | **Never** | **Inconsistent** | **Always** |  |
| **Without FBSW** | Negative | 23 169 (60.0%) | 7542 (19.5%) | 7888 (20.4%) | 38 599 |
|  | Positive | 4386 (60.0%) | 1554 (21.2%) | 1375 (18.8%) | 7315 |
| **With FBSW** | Negative | 1345 (38.1%) | 366 (10.4%) | 1821 (51.6%) | 3532 |
|  | Positive | 900 (45.7%) | 220 (11.2%) | 848 (43.1%) | 1968 |

Abbreviations: FBSW, female bar or sex worker; HIV, human immunodeficiency virus
